# Clear Cell Renal Cell Carcinoma Metastases Visibility: Split-Bolus Single Scan vs. Portal Venous Phase CT

**DOI:** 10.1101/2025.08.09.25333372

**Authors:** Bulent Aslan, Francesca Rigiroli, Alexander Brook, Olga Rachel Brook

## Abstract

**Objective:** To assess the impact of split-bolus (SB) single scan CT on the conspicuity of clear cell renal cell carcinoma (ccRCC) metastases compared to single-bolus injection.

**Methods:** This retrospective cohort study included consecutive patients with histologically proven metastatic ccRCC who underwent both SB and single-bolus portal venous abdominal CT within 6 months between 2017 and 2022 in a single tertiary center. SB CT utilized BMI-adjusted contrast dose and kVp (80-120) with concurrent arterial and portal venous phases. Single bolus CT utilized BMI-adjusted contrast dose at 120kVp at the portal venous phase. Wilcoxon rank test compared the conspicuity of metastases between the protocols.

**Results:** Of the 47 patients, 80.9% were male, with a mean age of 67 ± 10.4 years and a BMI of 27.1 ± 5.7. There were 48 paired CTs performed with a median interval of 93 days. Contrast dose was 143 ± 27 cc for SB and 108 ± 26 cc for single-bolus (*p*< 0.001). 66 metastases were analyzed, with an average size of 2 cm: 48.5% in the pancreas, 28.8% in skeletal muscle, and 22.7% in the liver. The median contrast-to-noise ratio (CNR) was higher with SB compared to single-bolus for all metastases (3.0 vs. 1.4), pancreatic metastases (2.7 vs. 1.4), muscle metastases (5.2 vs. 2.0), and liver metastases (2.8 vs. 0.9), all *p*< 0.001.

**Conclusions:** SB scan provides dramatically higher conspicuity of ccRCC metastases as compared to single-bolus portal venous CT.

## Introduction

Clear cell renal cell carcinoma (ccRCC) is the most common histologic subtype of renal cell carcinoma (RCC)[1]. It represents approximately 75-80% of all RCC cases and frequently has metastatic disease[2]. The most common metastatic sites of ccRCC are lungs and bones[3]. Some of other less common but important sites of metastases are liver, pancreas, and soft tissues[3]. The clear cell histologic subtype of renal carcinoma represents a notably hypervascular tumor characterized by an extensive network of blood vessels[4], which is also present in its metastases[5].

Routine oncological imaging is usually performed in portal venous phase, as this is an optimal phase for visceral organs evaluation of most common hypovascular metastases[6]. However, in case of RCC due to its hypervascularity, late arterial phase is crucial for detection and evaluation for both primary lesion and metastatic sites[7]. Current Society of Abdominal Radiology RCC Disease-Focused Panel RCC imaging recommendation include CT in the portal venous phase (60–90 seconds), with optional imaging of the abdomen in the late arterial phase (40–50 seconds) to help improve detection of hypervascular metastases in the liver and pancreas[8]. Dual phase protocol results in double number of images to review and increased radiation exposure, as opposed to standard portal venous phase oncological imaging. While detection of RCC metastases is possible in portal venous phase, it is challenging with decreased lesions conspicuity at the portal venous phase[9].

We developed an alternative protocol that uses two contrast injections with a single scan to produce simultaneous arterial and portal venous phases in one scan. The protocol utilizes BMI-adjusted contrast dose and lower kVp in smaller patients to further improve attenuation of enhancing lesions. Split bolus single scan technique has been shown to provide good quality simultaneous arterial and portal venous phase imaging for clinical indications where both phases are important for detection and characterization of lesions, such as CT enterography, mesenteric ischemia, pancreatic cancer staging[10–12]. We hypothesize that the split bolus single scan study can provide higher conspicuity of the ccRCC metastases with similar radiation dose and number of images as in portal venous phase study.

The goal of this study was to assess the impact of split-bolus (SB) single scan CT on the conspicuity of metastatic lesions compared to single-bolus injection in the same patients with clear cell renal cell carcinoma (ccRCC), evaluating the same metastatic lesions in both techniques.

## Materials and Methods

### Institutional Review Board

This retrospective study was conducted with the approval of the institutional review board and was compliant with the Health Insurance Portability and Accountability Act regulations. The requirement for informed consent was waived by the institutional review board due to retrospective nature of the study.

### Study design

The study population comprised of retrospective consecutive cohort of patients with metastatic renal cell carcinoma that underwent CT studies with split bolus and general oncology protocol from January 1, 2017, to December 31, 2022, with time interval of less than 6 months between the studies. This enabled a paired design that allows to compare conspicuity of metastases in the liver, pancreas, and skeletal muscle with two protocols. The time interval of six months ensures that no significant changes occurred in the interim in enhancement of the lesions. Patients were excluded from the study if they had non-clear cell histology, evidence of treated metastases, or a time interval greater than 6 months between the single-bolus and split-bolus CT scans (Figure 1).

**Figure 1:**
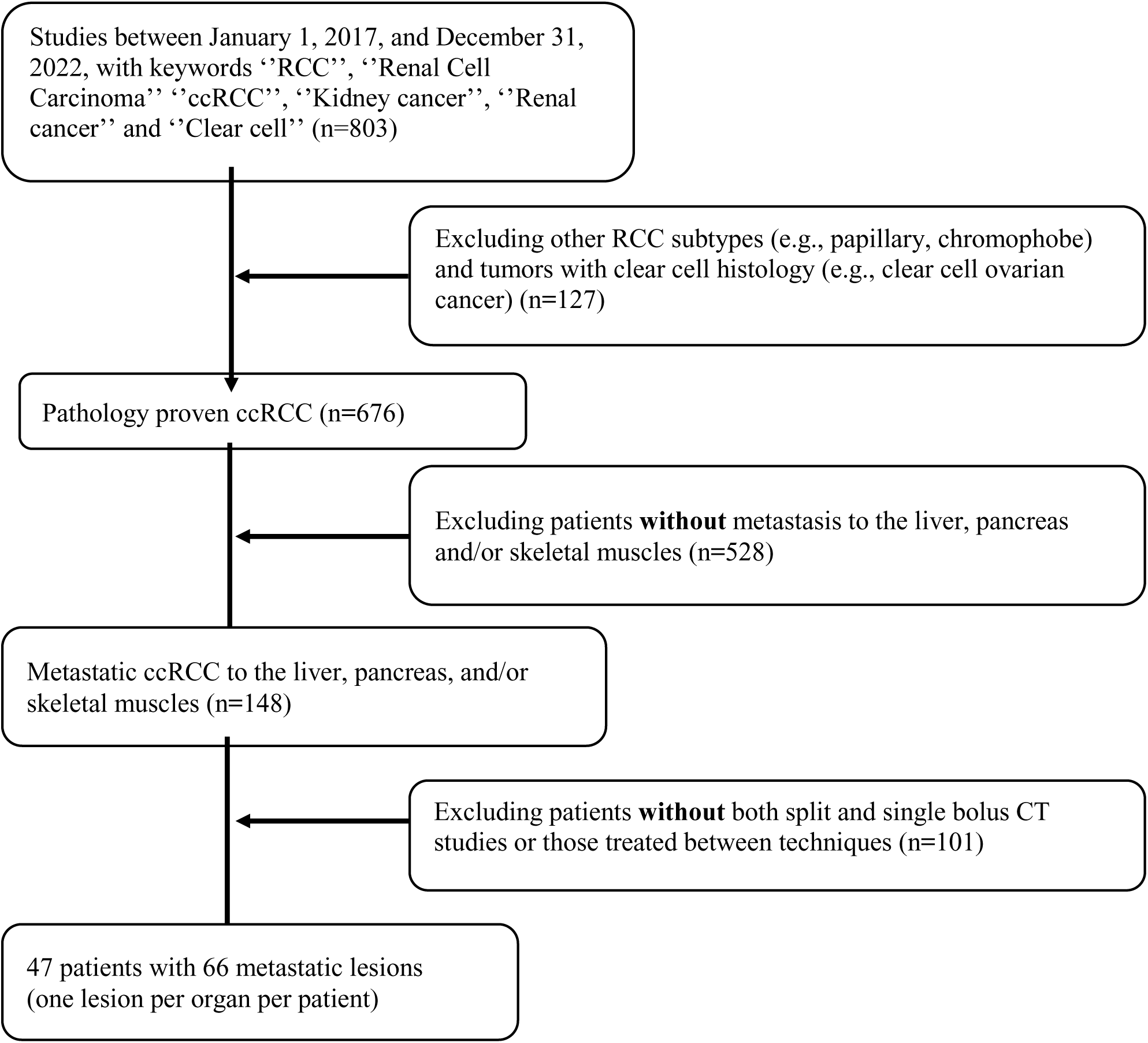
Patient selection.

The standard protocol in our institution for the follow up of patients with RCC is split-bolus CT abdomen and pelvis. However, at times, (when patients present for evaluation for other indications or wrong protocol is utilized for RCC follow up) patients are scanned with general oncology protocol, performed with portal venous phase. General oncology protocol is utilized for patients with non-RCC malignancies or non-specific symptoms. Therefore, some patients with metastatic RCC had been scanned with both protocols within short time period. The patients were identified through a search of institutional CT database for reports of abdomen and pelvis CT studies that contain terms such as renal cell carcinoma, RCC, ccRCC, kidney cancer, renal cancer, kidney carcinoma or renal carcinoma. All patients included in the study had histologically proven ccRCC with metastatic spread that included the liver, pancreas, and skeletal muscles. Metastasis was proven either with histology or follow-up of at least 2 years.

### Data Analysis

Conspicuity assessment of the lesions was performed by a clinical abdominal radiology fellow (X). A subset of random lesions (n=15) was also remeasured by additional clinical abdominal radiology fellow (Y) to assess interobserver variability for measurement accuracy. A maximum of one metastatic lesion per organ per patient was included.

The following data were collected for all patients: age, gender, height, and weight as documented in the online medical records. Body mass index (BMI) was calculated as kilograms divided by the square of height in meters. CT radiation exposure data including volumetric CT dose index (CTDI) and dose length product (DLP) were obtained from the picture archiving and communications systems (PACS) where it is reported automatically from the scanner data.

### Scanning Parameters

CT scans were performed in single energy mode on three different MDCT scanners: 64-MDCT Discovery HD (GE Healthcare), 128-MDCT Definition AS (Siemens Healthcare), and 320-MDCT Aquilion One (Toshiba Healthcare). 3D automatic exposure control was utilized in all scans. Axial, sagittal, and coronal reconstructions were performed with a medium level of iterative reconstruction (ADIR SD 12.5, Toshiba Healthcare; 30% ASIR, GE Healthcare; SAFIRE Level 2, Siemens Healthcare).

### Split-bolus single scan CT technique

Bolus-triggering was used to start scan with a threshold of 250–360 HU in the abdominal aorta at the level of the diaphragm according to kVp (80 kVp - 360 HU, 100 kVp - 300 HU and 120 kVp - 250 HU). A total volume of 90-110 mL of contrast agent (Omnipaque 350, GE Healthcare) depending on the patient’s BMI was injected IV with an injection rate of 2-4.5 mL/s using a power injector. The contrast agent was divided into an initial 40-60 mL bolus, followed by a 23-38 second delay and a second bolus of 50 mL of contrast agent. A 40 mL saline chaser followed the contrast injection. The tube voltage was selected by the technologist at the time of the study in split-bolus injection technique according to the patient’s BMI: 80 kVp for patients with BMI less than 20, 100 kVp for BMI of 20–30, and 120 kVp for BMI greater than 30 (Table 1).

**Table 1:** Technical Parameters of the Single-Bolus Injection and Split-Bolus Injection CT Abdomen and Pelvis Protocols.

| **Parameter** | **Single-Bolus portal venous** | **Split-Bolus single scan combining arterial and portal venous phases** |
| --- | --- | --- |
| Bolus triggering | Trigger set 50 HU above liver inherent HU | 80 kVp: 360 HU<br>100 kVp: 300 HU<br>120 kVp: 250 HU |
| Tube voltage | 120 kVp | BMI < 20: 80 kVp<br>BMI 20-30: 100 kVp<br>BMI > 30: 120 kVp |
| Maximum tube current (mA) | 600 | 600 |
| Slice thickness (mm) | 2.5 | 2.5 |
| Pitch | 0.984 | 0.984 |
| Collimation (mm) | 0.625 | 0.625 |
| Contrast amount (mL) | BMI < 20: 60 mL<br>BMI 20-30: 80 mL<br>BMI > 30: 100 mL | BMI < 20: 90 mL<br>BMI 20-30: 100 mL<br>BMI > 30: 110 mL |

### Standard single-bolus CT technique

CT scan was started using a bolus-triggering technique after the threshold of 50 HU above the liver inherent HU was reached in the abdominal aorta at the level of the diaphragm. A total volume of 60-100 mL of contrast agent (Omnipaque 350, GE Healthcare) according to patient’s BMI was injected at an injection rate of 1.9-4 mL/s using a power injector. The contrast agent was injected as a single bolus followed by a 30-mL saline chaser. The tube voltage was selected as 120 kVp for all patients in standard single-bolus injection technique (Table 1).

### Objective Image Analysis

Following measurements at standard portal venous phase and split bolus scan were obtained by clinical fellow (X): mean attenuation of the aorta at the level of celiac trunk, main portal vein, metastatic lesions, background normal organ or tissue; standard deviation (SD) of the air, metastatic lesions and background normal organ or tissue. Two regions of interest (ROI) were taken in each location, and the average of these was recorded as a final measurement. The contrast-to-noise ratio (CNR) was calculated for each metastatic lesion using the formula.

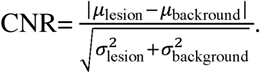

Here μlesion represents the mean intensity or value of the pixels within the lesion or region of interest, μbackground represents the mean intensity or value of the pixels in the background or surrounding area, σlesion is the standard deviation of the pixel values within the lesion, σbackground is the standard deviation of the pixel values in the background or surrounding area.

### Statistical Analysis

Statistical evaluation was performed using Matlab v.9. Descriptive statistics were calculated for patient demographics, including means and standard deviations for continuous variables such as age, weight, height, and BMI, and frequencies with percentages for categorical variables. The primary outcome measure, contrast-to-noise ratio (CNR) of metastatic lesions, was compared between split-bolus and single-bolus CT scans using the Wilcoxon signed-rank test due to the paired nature of the data.

Differences in lesion size, CT radiation exposure parameters (CTDI and DLP), and contrast volumes were also assessed with the Wilcoxon test. Interobserver variability in CNR measurements was determined using the Pearson correlation. A two-tailed p-value of less than 0.05 was considered statistically significant.

## Results

There were 47 patients included in the study. There were 38/47 (80.9%) male, mean age of 67 ± 10.4 years, mean BMI of 27.1 ± 5.7. There were 48 paired CT scans performed with split-bolus single scan and standard single-bolus scan techniques. The contrast dose was higher for split bolus studies with 143 ± 27 cc vs. 108 ± 26 cc in single bolus studies, *p* < 0.001. Sixty-six metastases were analyzed, with the median size of 1.6 cm [IQR 1 -2.6]) in single bolus technique and 1.8 cm in split bolus technique [IQR 1.2 – 2.7]), 32/66 (48.5%) were located in the pancreas, 19/66 (28.8%) in skeletal muscle, and 15/66 (22.7%) in the liver. (Table 2).

**Table 2:** Characteristics of ccRCC Patients with Metastasis to Liver, Pancreas or Skeletal Muscles Age 67 ± 10.4.

|  |  |
| --- | --- |
| Age | 67 ± 10.4 |
| Gender | female 9 (19.2%)<br>male 38 (80.9%) |
| Weight (kg) | 81.1 ± 18.7 |
| Height (cm) | 173 ± 11.4 |
| BMI | 27.1 ± 5.73 |
| Metastatic sites | Pancreas: 32 (48.48%)<br>Liver: 15 (22.73%)<br>Skeletal muscle: 19 (28.79%) |

The average interval between single bolus vs split bolus injection CT studies was 110 ± 66 days, median of 93 days [IQR 64.5-138]. In 37/48 (77%) paired CT studies, the split-bolus technique was performed after a single-bolus study.

There was higher contrast amount administered for split bolus studies as compared to single-bolus method (average 143 ml vs. 108 ml, *p* < 0.001, median 150 ml [IQR 120 -160] vs. 100 ml ([IQR 100 - 130]). There was no notable difference in Computed Tomography Dose Index (CTDI) between the split and single bolus techniques (avg=14.5 ± 5.36, med=13.2 [IQR 9.9 -18.05] and avg=15.3 ± 7.65, med=12.7 [IQR 10.4-18.6] respectively, *p* = 0.7). Background noise (air SD) also displayed no significant variance between split-bolus and single-bolus protocols (Table 3).

**Table 3:** Imaging Characteristics of ccRCC Patients with Metastasis to Liver, Pancreas or Skeletal Muscle.

|  | Single Bolus Injection | Split Bolus Injection | p-value |
| --- | --- | --- | --- |
| CTDI (mGy) | 15.3 ± 7.7 | 14.5 ± 5.4 | 0.67 |
| Background Noise (Air SD) | 7.1 ± 2.2 | 7.4 ± 1.7 | 0.4 |
| Lesion size (cm) | 2.0 ± 1.4 | 2.1 ± 1.3 | 0.023 |
| Aortic Enhancement (HU) | 162 ± 62 | 315 ± 138 | < 0.001 |
| Portal Vein Enhancement (HU) | 160 ± 40 | 211 ± 54 | < 0.001 |
| CNR all sites, median [IQR] | 1.4 [0.8–2.3] | 3.0 [2.3–5.4] | < 0.001 |
| Liver metastases | 0.9 [0.6–1.4] | 2.8 [2.1–4.0] | < 0.001 |
| Pancreatic metastases | 1.4 [0.8–2.0] | 2.7 [2.3–5.1] | < 0.001 |
| Skeletal muscle metastases | 2.0 [1.4–3.1] | 5.2 [3.0–7.6] | < 0.001 |

### Lesion Conspicuity

The split-bolus method demonstrated higher attenuation of the aorta (median 276 [IQR 219 -371] vs. 145 [IQR 129 -168]) and portal vein (median=202 [IQR 184 -241] vs. 168 [IQR 125 -186]) compared to single-bolus CT, (*p* < 0.001 for both comparisons).

The median contrast-to-noise ratio (CNR) was higher with split bolus compared to single-bolus for all metastases (3.0 [IQR 2.3-5.4] vs. 1.4 [IQR 0.8-2.3]). Similar results were seen for pancreatic metastases (2.7 [IQR 2.3-5.1] vs. 1.4 [IQR 0.8-2.0]), muscle metastases (5.2 [IQR 3.0-7.6] vs. 2.0 [IQR 1.4-3.1]), and liver metastases (2.8 [IQR 2.1-4.0] vs. 0.9 [IQR 0.6-1.4]), all *p* < 0.001 (Figures 2 and 3).

**Figure 2:**
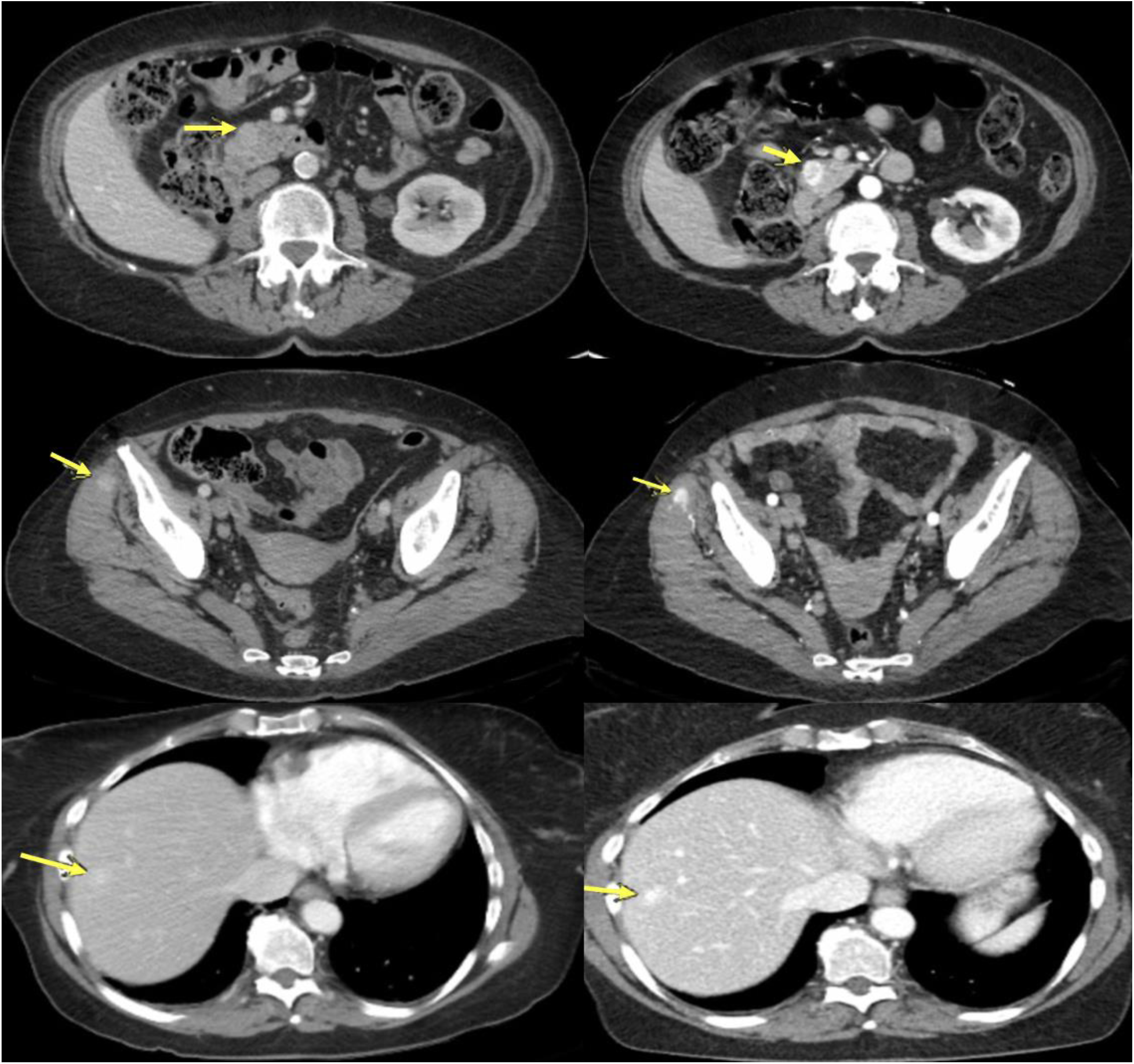
Single Bolus vs Split Bolus CT scans for various sites of metastases. Axial CT images of the pancreas (upper row), skeletal muscle (middle row), and liver (lower row) metastases in single bolus (left) and split bolus (right) techniques in 3 different patients with metastatic RCC. Yellow arrows indicate metastases.

**Figure 3:**
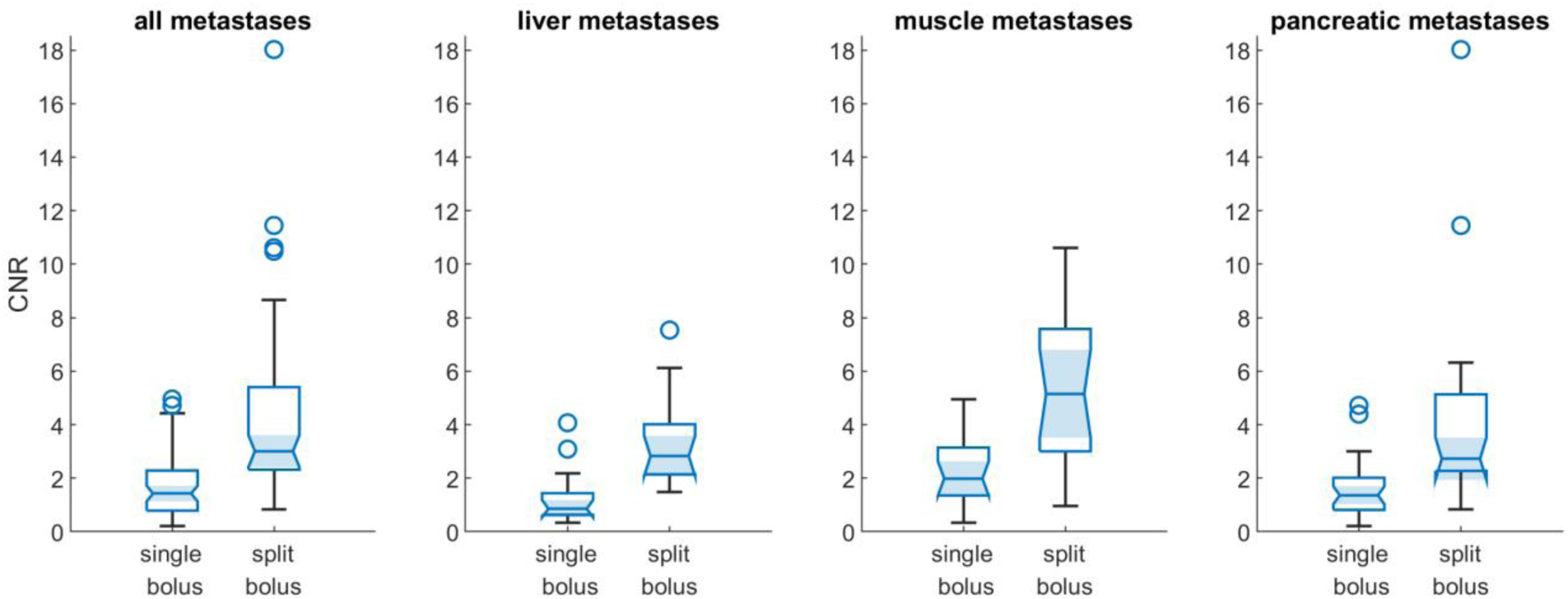
Comparison of the CNR of the metastatic lesions in the single-bolus vs split-bolus injection techniques.

### Validation of measurement by assessment of interobserver variability

We assessed validity of a single investigator measuring density of the lesions with both techniques by assessing for interobserver variability on a subset of 15 patients. There was strong correlation in lesion and background density measurements with correlation coefficients ranging between 0.98 and 0.99 for both split bolus and single bolus techniques.

## Discussion

Our results show the effectiveness of split-bolus single scan CT with contrast dose and voltage BMI-based adjustments in enhancing detection and visualization of metastatic lesions in patients with clear cell renal cell carcinoma (ccRCC) when compared to the conventional single-bolus injection method. There was a significantly improved lesion conspicuity in all sites of metastases with split bolus protocol, while keeping radiation dose and noise level similar to the standard protocol. Overall, these findings demonstrate the potential of split-bolus single scan CT as a valuable tool in optimizing diagnostic accuracy for ccRCC patients by enhancing lesion visualization and delineation.

Significant increase in contrast-to-noise ratio (CNR) of ccRCC metastases in the proposed protocol utilizing split-bolus injection technique compared to the single-bolus method is likely multifactorial. First, the multiphasic scan, providing simultaneous arterial and portal venous phases increases attenuation of arterially enhancing metastases[13]. Second, the amount of contrast is mildly increased. Third, we utilized lower kVp in smaller patients, which increases attenuation of iodine and thus lesional enhancement[14]. Tailored protocols for specific indications can be difficult to implement consistently in busy clinical centers, however, as shown in these results, they provide better patient care than “one-fit-all” protocol. Automation and auditing are key components in successful patient-specific medicine and protocol selection is amenable for quality improvement methodology.

Our research extends the application of the split-bolus injection technique to evaluating tumor metastases in clear cell renal cell carcinoma (ccRCC). Building on prior studies showing improved detection of mucosal hyperenhancement in inflammatory bowel disease, enhanced tumor conspicuity in pancreatic imaging, and confident diagnostic ability in assessing mesenteric ischemia, we explored this technique for detection of metastatic ccRCC lesions[10–12]. Consistent with previous findings, the split-bolus approach demonstrated enhanced lesion visualization while potentially reducing radiation exposure. By validating this technique for ccRCC metastases, our study adds to the growing evidence supporting the split-bolus technique’s efficacy in improving diagnostic accuracy while minimizing radiation dose and interpretation time.

The slightly larger size of metastatic lesions observed in the split-bolus injection scans compared to the single-bolus scans can be attributed to either the time interval between the two imaging acquisitions or “blooming” of the lesion due to its increased attenuation[15]. As the split-bolus scans were predominantly performed after the single-bolus scans, the intervening time period may have allowed for lesion growth or progression, resulting in the increased dimensions captured during the split-bolus imaging. The difference was not substantial (1mm), therefore not likely of clinical significance.

Radiation exposure, as demonstrated by CTDI index, was similar with split bolus and single bolus protocols. Therefore, increased lesion conspicuity with split bolus protocol was obtained without increase radiation dose. Patients with RCC are usually imaged for many years due to somewhat indolent behavior of the tumor, therefore decreasing overall radiation exposure in these patients is an important consideration[16,17].

Another critical aspect of the split-bolus injection technique’s success also potentially lies in the adjustment of kilovoltage peak (kVp) according to patient BMI. Lowering the kVp significantly contributes to increased attenuation of iodine-containing structures by bringing it closer to the k-edge of iodine[18]. This principle is effectively utilized in CT angiography (CTA) to enhance vascular contrast[19]. However, it is essential to acknowledge the trade-off involved: while lower kVp imaging increases iodine attenuation and subsequently improves lesion conspicuity, it also results in higher image noise. This inherent increase in noise is why low kVp imaging is not routinely employed across all CT protocols[20]. Despite this, our study demonstrates that the benefits of using a BMI-based kVp adjustment, specifically in the context of ccRCC metastases detection, outweigh the drawbacks by substantially enhancing the contrast-to-noise ratio without a significant rise in overall radiation dose. This tailored approach exemplifies how protocol adjustments can be optimized to leverage the physical properties of contrast media, ultimately improving diagnostic accuracy in oncologic imaging.

The substantially higher CNR not only facilitates superior lesion delineation but may also potentially contribute to improved diagnostic confidence and accuracy, particularly in identifying smaller or inconspicuous lesions that may otherwise be overlooked or challenging to detect, especially for less experienced radiologists and in high-volume imaging centers. Translation of lesional conspicuity clinical impact is not a given. Hagen et al showed no benefit in accuracy of dual phase CT as compared to single portal venous imaging for patients with renal cell carcinoma[21]. However, that study include patients with metastases in organs with high inherent CNR, such as lung and adrenal, while our study focused on patients with metastases to the organs with lower inherent CNR. Clinical impact (beyond improved conspicuity) of the proposed split bolus protocol on detection of metastases should be explored in the future studies.

This study has several limitations that warrant consideration. First, the retrospective nature introduces inherent biases and constraints associated with data collection and analysis from pre-existing institutional databases. Another key limitation is the time interval between the single-bolus and split-bolus scans, where observed differences in lesion size could partly reflect disease progression during this period rather than solely the imaging technique’s performance. Furthermore, our study evaluates imaging parameters but lacks direct clinical outcome measures or long-term follow-up data, necessitating further research to establish the enhanced lesion conspicuity’s direct impact on patient management and outcomes. Additionally, while the split-bolus technique demonstrated improved lesion visualization compared to single-bolus imaging, the study did not directly compare it with the dual-phase (arterial plus portal venous) protocol, which could have provided a more comprehensive assessment of its relative efficacy. Prospective studies with direct comparisons across imaging protocols and incorporation of clinical outcomes are needed to further validate the split-bolus technique’s utility in this clinical context.

In conclusion, our study supports the split-bolus injection technique with BMI-adjusted contrast dose and scanning parameters as a promising approach for improving metastatic lesion detection in ccRCC patients, demonstrated by the notable increase in contrast-to-noise ratio and enhanced lesion conspicuity.

## Data Availability

All data produced in the present study are available upon reasonable request to the authors

